## Supplementary material for "A molnupiravir-associated mutational signature in global SARS-CoV-2 genomes": GISAID Acknowledgements 1

### SUPPLEMENTAL TABLE

**Data Availability**

GISAID Identifier: EPI_SET_230110wz
doi: 10.55876/gis8.230110wz

All genome sequences and associated metadata in this dataset are published in GISAID's EpiCoV database. To view the contributors of each individual sequence with details such as accession number, Virus name, Collection date, Originating Lab and Submitting Lab and the list of Authors, visit 10.55876/gis8.230110wz

**Data Snapshot**

- EPI_SET_230110wz is composed of 7,748,305 individual genome sequences.
- The collection dates range from 2020-01-01 to 2022-12-29;
- Data were collected in 213 countries and territories;
- All sequences in this dataset are compared relative to hCoV-19/Wuhan/WIV04/2019 (WIV04), the official reference sequence employed by GISAID (EPI_ISL_402124). Learn more at https://gisaid.org/WIV04.
