## Appendix of cluster trees for "A molnupiravir-associated mutational signature in global SARS-CoV-2 genomes"

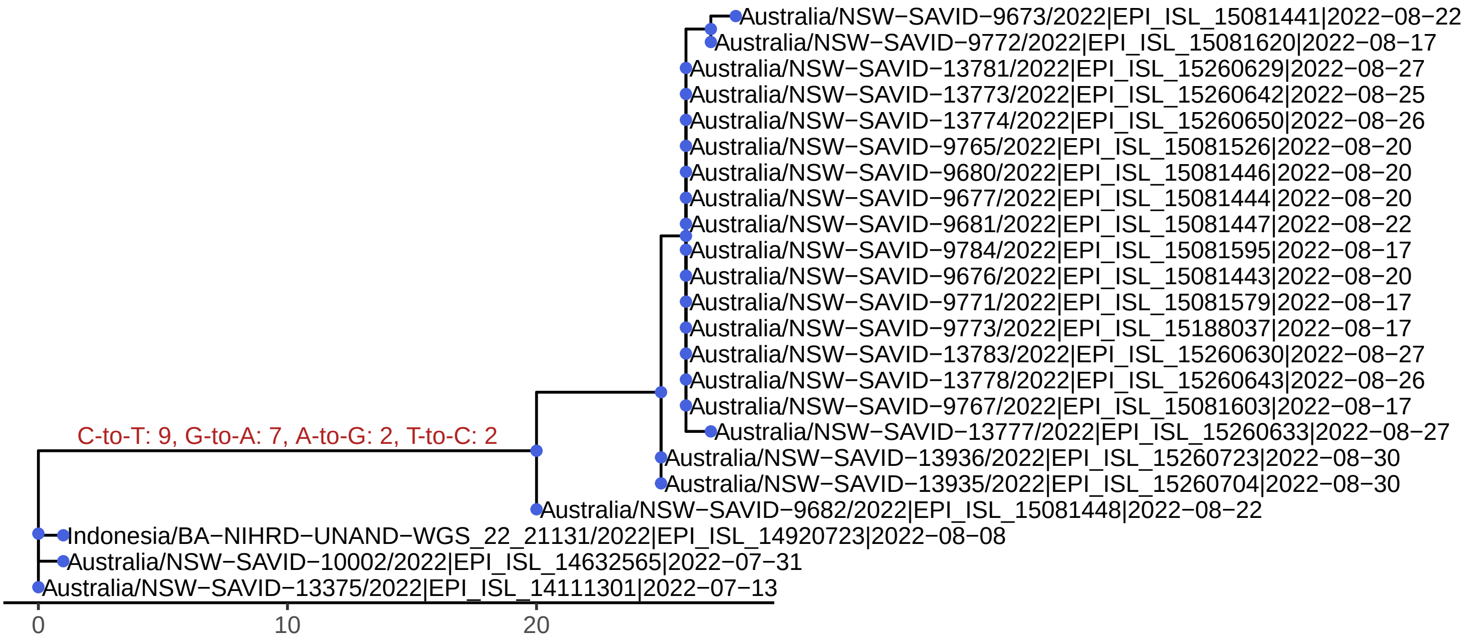
Australia/NSW−SAVID−9673/2022|EPI_ISL_15081441|2022−08−22
Australia/NSW−SAVID−9772/2022|EPI_ISL_15081620|2022−08−17
Australia/NSW−SAVID−13781/2022|EPI_ISL_15260629|2022−08−27
Australia/NSW−SAVID−13773/2022|EPI_ISL_15260642|2022−08−25
Australia/NSW−SAVID−13774/2022|EPI_ISL_15260650|2022−08−26
Australia/NSW−SAVID−9765/2022|EPI_ISL_15081526|2022−08−20
Australia/NSW−SAVID−9680/2022|EPI_ISL_15081446|2022−08−20
Australia/NSW−SAVID−9677/2022|EPI_ISL_15081444|2022−08−20
Australia/NSW−SAVID−9681/2022|EPI_ISL_15081447|2022−08−22
Australia/NSW−SAVID−9784/2022|EPI_ISL_15081595|2022−08−17
Australia/NSW−SAVID−9676/2022|EPI_ISL_15081443|2022−08−20
Australia/NSW−SAVID−9771/2022|EPI_ISL_15081579|2022−08−17
Australia/NSW−SAVID−9773/2022|EPI_ISL_15188037|2022−08−17
Australia/NSW−SAVID−13783/2022|EPI_ISL_15260630|2022−08−27
Australia/NSW−SAVID−13778/2022|EPI_ISL_15260643|2022−08−26
Australia/NSW−SAVID−9767/2022|EPI_ISL_15081603|2022−08−17
C-to-T: 9, G-to-A: 7, A-to-G: 2, T-to-C: 2
Australia/NSW−SAVID−13777/2022|EPI_ISL_15260633|2022−08−27
Australia/NSW−SAVID−13936/2022|EPI_ISL_15260723|2022−08−30
Australia/NSW−SAVID−13935/2022|EPI_ISL_15260704|2022−08−30
Australia/NSW−SAVID−9682/2022|EPI_ISL_15081448|2022−08−22
Indonesia/BA−NIHRD−UNAND−WGS_22_21131/2022|EPI_ISL_14920723|2022−08−08
Australia/NSW−SAVID−10002/2022|EPI_ISL_14632565|2022−07−31
Australia/NSW−SAVID−13375/2022|EPI_ISL_14111301|2022−07−13
0
10
20

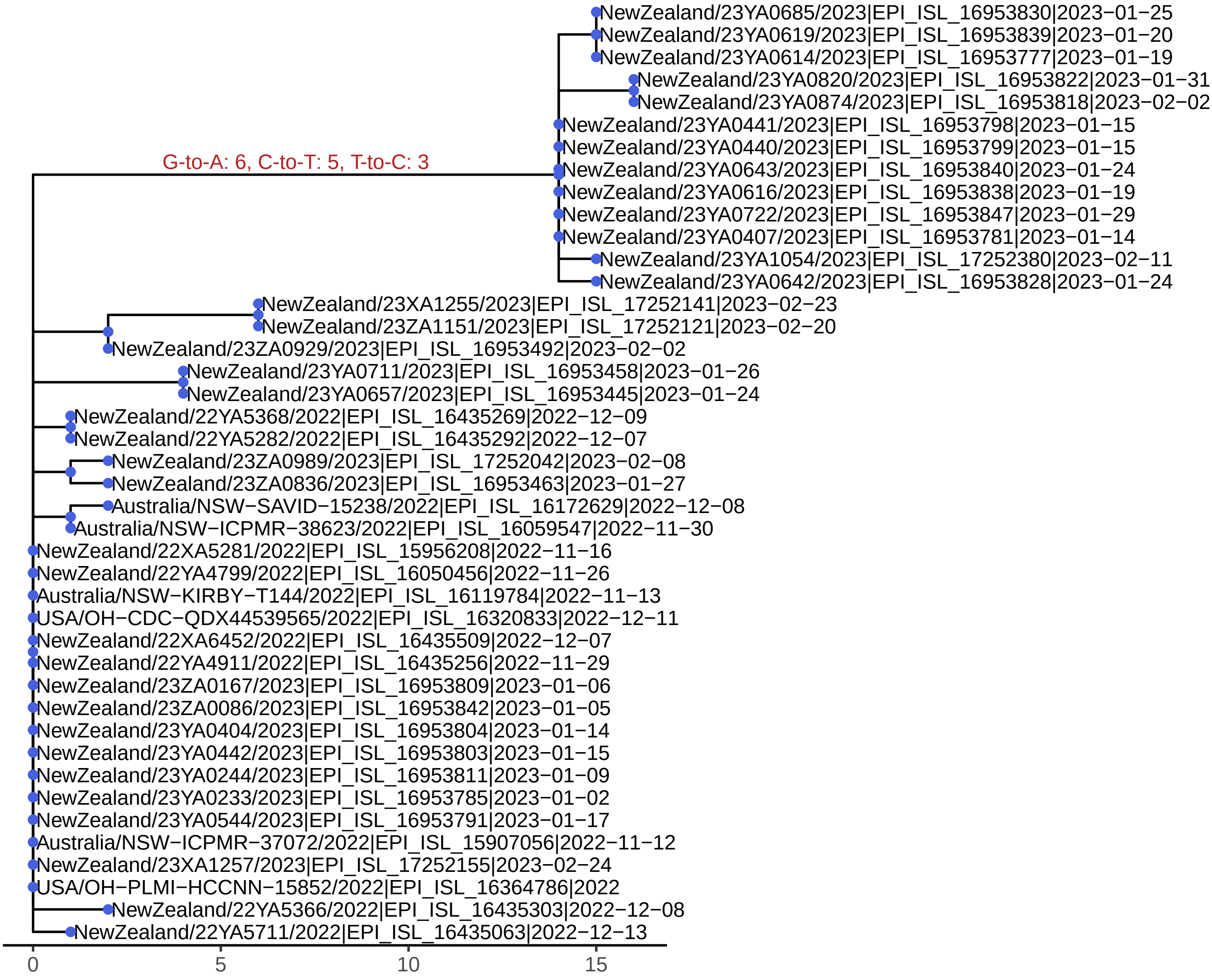
NewZealand/23YA0685/2023|EPI_ISL_16953830|2023−01−25
NewZealand/23YA0619/2023|EPI_ISL_16953839|2023−01−20
NewZealand/23YA0614/2023|EPI_ISL_16953777|2023−01−19
NewZealand/23YA0820/2023|EPI_ISL_16953822|2023−01−31
NewZealand/23YA0874/2023|EPI_ISL_16953818|2023−02−02
NewZealand/23YA0441/2023|EPI_ISL_16953798|2023−01−15
NewZealand/23YA0440/2023|EPI_ISL_16953799|2023−01−15
G-to-A: 6, C-to-T: 5, T-to-C: 3
NewZealand/23YA0643/2023|EPI_ISL_16953840|2023−01−24
NewZealand/23YA0616/2023|EPI_ISL_16953838|2023−01−19
NewZealand/23YA0722/2023|EPI_ISL_16953847|2023−01−29
NewZealand/23YA0407/2023|EPI_ISL_16953781|2023−01−14
NewZealand/23YA1054/2023|EPI_ISL_17252380|2023−02−11
NewZealand/23YA0642/2023|EPI_ISL_16953828|2023−01−24
NewZealand/23XA1255/2023|EPI_ISL_17252141|2023−02−23
NewZealand/23ZA1151/2023|EPI_ISL_17252121|2023−02−20
NewZealand/23ZA0929/2023|EPI_ISL_16953492|2023−02−02
NewZealand/23YA0711/2023|EPI_ISL_16953458|2023−01−26
NewZealand/23YA0657/2023|EPI_ISL_16953445|2023−01−24
NewZealand/22YA5368/2022|EPI_ISL_16435269|2022−12−09
NewZealand/22YA5282/2022|EPI_ISL_16435292|2022−12−07
NewZealand/23ZA0989/2023|EPI_ISL_17252042|2023−02−08
NewZealand/23ZA0836/2023|EPI_ISL_16953463|2023−01−27
Australia/NSW−SAVID−15238/2022|EPI_ISL_16172629|2022−12−08
Australia/NSW−ICPMR−38623/2022|EPI_ISL_16059547|2022−11−30
NewZealand/22XA5281/2022|EPI_ISL_15956208|2022−11−16
NewZealand/22YA4799/2022|EPI_ISL_16050456|2022−11−26
Australia/NSW−KIRBY−T144/2022|EPI_ISL_16119784|2022−11−13
USA/OH−CDC−QDX44539565/2022|EPI_ISL_16320833|2022−12−11
NewZealand/22XA6452/2022|EPI_ISL_16435509|2022−12−07
NewZealand/22YA4911/2022|EPI_ISL_16435256|2022−11−29
NewZealand/23ZA0167/2023|EPI_ISL_16953809|2023−01−06
NewZealand/23ZA0086/2023|EPI_ISL_16953842|2023−01−05
NewZealand/23YA0404/2023|EPI_ISL_16953804|2023−01−14
NewZealand/23YA0442/2023|EPI_ISL_16953803|2023−01−15
NewZealand/23YA0244/2023|EPI_ISL_16953811|2023−01−09
NewZealand/23YA0233/2023|EPI_ISL_16953785|2023−01−02
NewZealand/23YA0544/2023|EPI_ISL_16953791|2023−01−17
Australia/NSW−ICPMR−37072/2022|EPI_ISL_15907056|2022−11−12
NewZealand/23XA1257/2023|EPI_ISL_17252155|2023−02−24
USA/OH−PLMI−HCCNN−15852/2022|EPI_ISL_16364786|2022
NewZealand/22YA5366/2022|EPI_ISL_16435303|2022−12−08
NewZealand/22YA5711/2022|EPI_ISL_16435063|2022−12−13
0
5
10
15

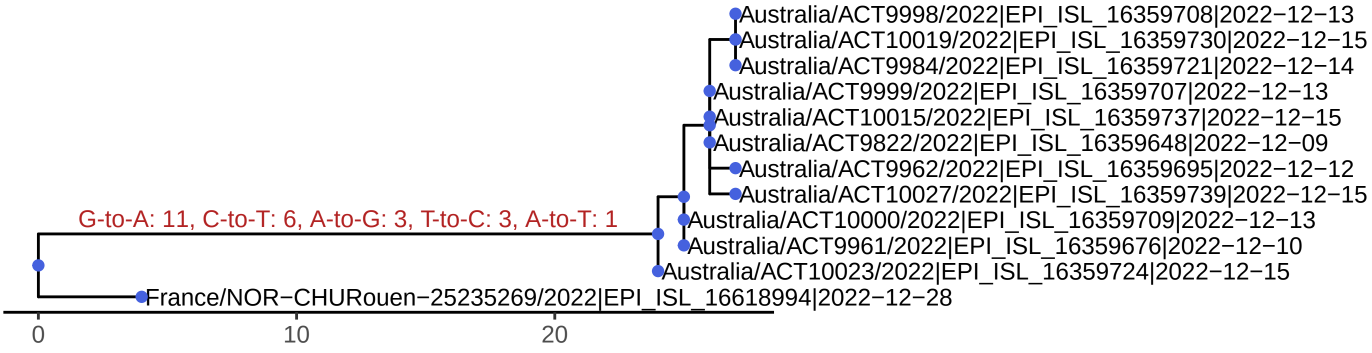
Australia/ACT9998/2022|EPI_ISL_16359708|2022−12−13
Australia/ACT10019/2022|EPI_ISL_16359730|2022−12−15
Australia/ACT9984/2022|EPI_ISL_16359721|2022−12−14
Australia/ACT9999/2022|EPI_ISL_16359707|2022−12−13
Australia/ACT10015/2022|EPI_ISL_16359737|2022−12−15
Australia/ACT9822/2022|EPI_ISL_16359648|2022−12−09
Australia/ACT9962/2022|EPI_ISL_16359695|2022−12−12
Australia/ACT10027/2022|EPI_ISL_16359739|2022−12−15
G-to-A: 11, C-to-T: 6, A-to-G: 3, T-to-C: 3, A-to-T: 1
Australia/ACT10000/2022|EPI_ISL_16359709|2022−12−13
Australia/ACT9961/2022|EPI_ISL_16359676|2022−12−10
Australia/ACT10023/2022|EPI_ISL_16359724|2022−12−15
France/NOR−CHURouen−25235269/2022|EPI_ISL_16618994|2022−12−28
0
10
20

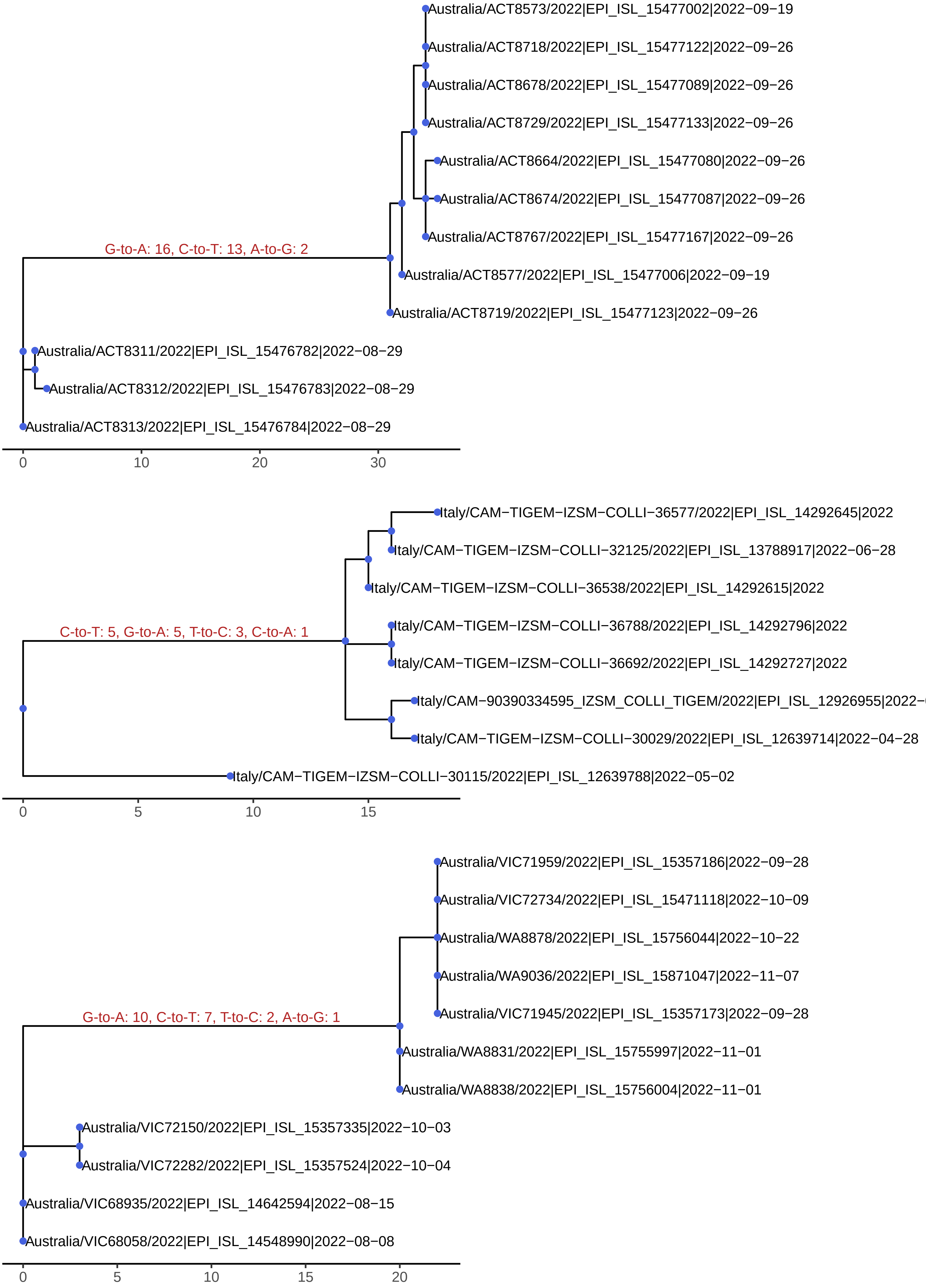
Australia/ACT8573/2022|EPI_ISL_15477002|2022−09−19
Australia/ACT8718/2022|EPI_ISL_15477122|2022−09−26
Australia/ACT8678/2022|EPI_ISL_15477089|2022−09−26
Australia/ACT8729/2022|EPI_ISL_15477133|2022−09−26
Australia/ACT8664/2022|EPI_ISL_15477080|2022−09−26
Australia/ACT8674/2022|EPI_ISL_15477087|2022−09−26
Australia/ACT8767/2022|EPI_ISL_15477167|2022−09−26
G-to-A: 16, C-to-T: 13, A-to-G: 2
Australia/ACT8577/2022|EPI_ISL_15477006|2022−09−19
Australia/ACT8719/2022|EPI_ISL_15477123|2022−09−26
Australia/ACT8311/2022|EPI_ISL_15476782|2022−08−29
Australia/ACT8312/2022|EPI_ISL_15476783|2022−08−29
Australia/ACT8313/2022|EPI_ISL_15476784|2022−08−29
0
10
20
30
Italy/CAM−TIGEM−IZSM−COLLI−36577/2022|EPI_ISL_14292645|2022
Italy/CAM−TIGEM−IZSM−COLLI−32125/2022|EPI_ISL_13788917|2022−06−28
Italy/CAM−TIGEM−IZSM−COLLI−36538/2022|EPI_ISL_14292615|2022
Italy/CAM−TIGEM−IZSM−COLLI−36788/2022|EPI_ISL_14292796|2022
C-to-T: 5, G-to-A: 5, T-to-C: 3, C-to-A: 1
Italy/CAM−TIGEM−IZSM−COLLI−36692/2022|EPI_ISL_14292727|2022
Italy/CAM−90390334595_IZSM_COLLI_TIGEM/2022|EPI_ISL_12926955|2022−
Italy/CAM−TIGEM−IZSM−COLLI−30029/2022|EPI_ISL_12639714|2022−04−28
Italy/CAM−TIGEM−IZSM−COLLI−30115/2022|EPI_ISL_12639788|2022−05−02
0
5
10
15
Australia/VIC71959/2022|EPI_ISL_15357186|2022−09−28
Australia/VIC72734/2022|EPI_ISL_15471118|2022−10−09
Australia/WA8878/2022|EPI_ISL_15756044|2022−10−22
Australia/WA9036/2022|EPI_ISL_15871047|2022−11−07
Australia/VIC71945/2022|EPI_ISL_15357173|2022−09−28
G-to-A: 10, C-to-T: 7, T-to-C: 2, A-to-G: 1
Australia/WA8831/2022|EPI_ISL_15755997|2022−11−01
Australia/WA8838/2022|EPI_ISL_15756004|2022−11−01
Australia/VIC72150/2022|EPI_ISL_15357335|2022−10−03
Australia/VIC72282/2022|EPI_ISL_15357524|2022−10−04
Australia/VIC68935/2022|EPI_ISL_14642594|2022−08−15
Australia/VIC68058/2022|EPI_ISL_14548990|2022−08−08
0
5
10
15
20

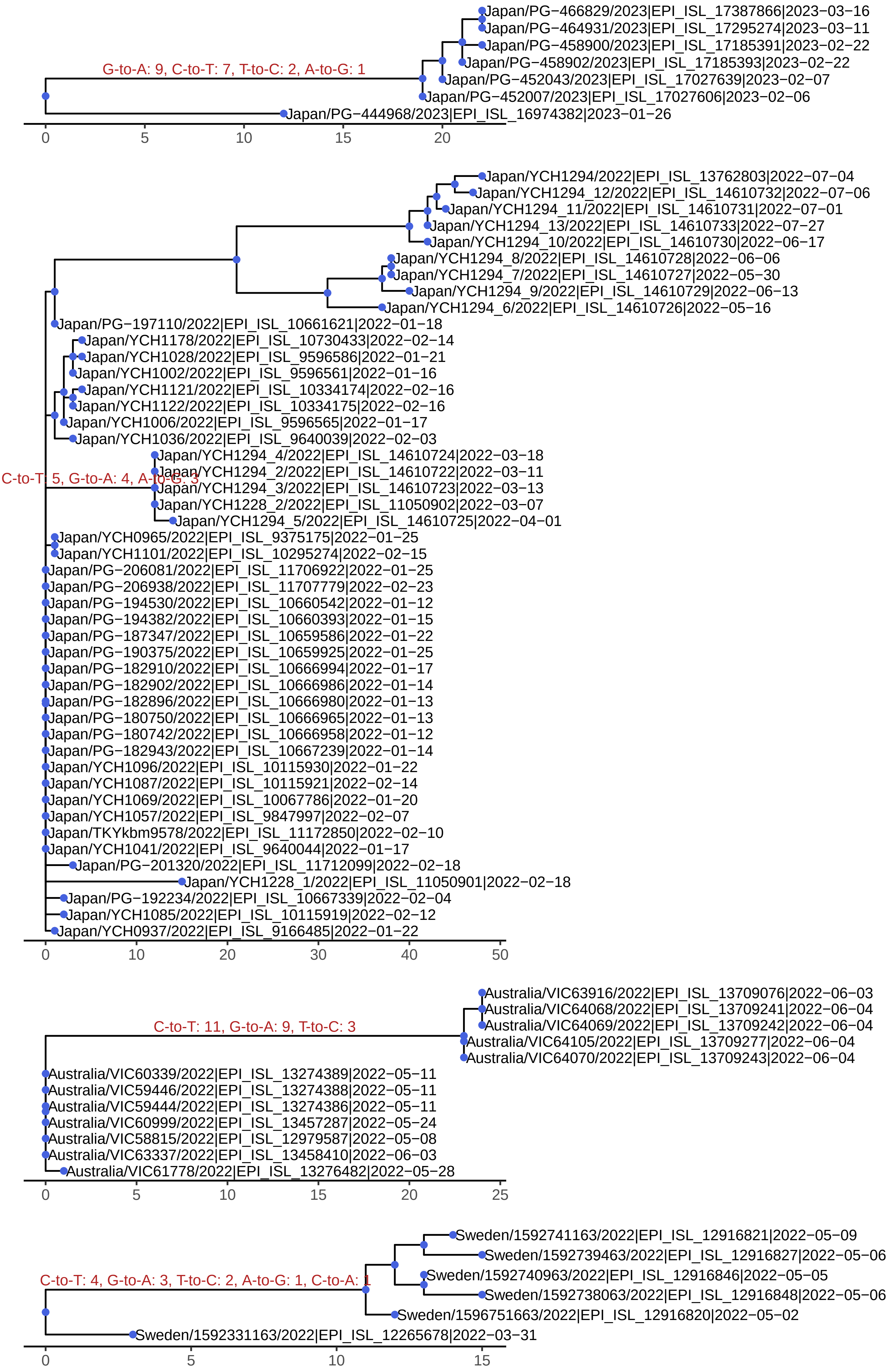
G-to-A: 9, C-to-T: 7, T-to-C: 2, A-to-G: 1
Japan/PG−466829/2023|EPI_ISL_17387866|2023−03−16
Japan/PG−464931/2023|EPI_ISL_17295274|2023−03−11
Japan/PG−458900/2023|EPI_ISL_17185391|2023−02−22
Japan/PG−458902/2023|EPI_ISL_17185393|2023−02−22
Japan/PG−452043/2023|EPI_ISL_17027639|2023−02−07
Japan/PG−452007/2023|EPI_ISL_17027606|2023−02−06
Japan/PG−444968/2023|EPI_ISL_16974382|2023−01−26
0
5
10
15
20
Japan/YCH1294/2022|EPI_ISL_13762803|2022−07−04
Japan/YCH1294_12/2022|EPI_ISL_14610732|2022−07−06
Japan/YCH1294_11/2022|EPI_ISL_14610731|2022−07−01
Japan/YCH1294_13/2022|EPI_ISL_14610733|2022−07−27
Japan/YCH1294_10/2022|EPI_ISL_14610730|2022−06−17
Japan/YCH1294_8/2022|EPI_ISL_14610728|2022−06−06
Japan/YCH1294_7/2022|EPI_ISL_14610727|2022−05−30
Japan/YCH1294_9/2022|EPI_ISL_14610729|2022−06−13
Japan/YCH1294_6/2022|EPI_ISL_14610726|2022−05−16
Japan/PG−197110/2022|EPI_ISL_10661621|2022−01−18
Japan/YCH1178/2022|EPI_ISL_10730433|2022−02−14
Japan/YCH1028/2022|EPI_ISL_9596586|2022−01−21
Japan/YCH1002/2022|EPI_ISL_9596561|2022−01−16
Japan/YCH1121/2022|EPI_ISL_10334174|2022−02−16
Japan/YCH1122/2022|EPI_ISL_10334175|2022−02−16
Japan/YCH1006/2022|EPI_ISL_9596565|2022−01−17
Japan/YCH1036/2022|EPI_ISL_9640039|2022−02−03
Japan/YCH1294_4/2022|EPI_ISL_14610724|2022−03−18
C-to-T: 5, G-to-A: 4, A-to-G: 3
Japan/YCH1294_2/2022|EPI_ISL_14610722|2022−03−11
Japan/YCH1294_3/2022|EPI_ISL_14610723|2022−03−13
Japan/YCH1228_2/2022|EPI_ISL_11050902|2022−03−07
Japan/YCH1294_5/2022|EPI_ISL_14610725|2022−04−01
Japan/YCH0965/2022|EPI_ISL_9375175|2022−01−25
Japan/YCH1101/2022|EPI_ISL_10295274|2022−02−15
Japan/PG−206081/2022|EPI_ISL_11706922|2022−01−25
Japan/PG−206938/2022|EPI_ISL_11707779|2022−02−23
Japan/PG−194530/2022|EPI_ISL_10660542|2022−01−12
Japan/PG−194382/2022|EPI_ISL_10660393|2022−01−15
Japan/PG−187347/2022|EPI_ISL_10659586|2022−01−22
Japan/PG−190375/2022|EPI_ISL_10659925|2022−01−25
Japan/PG−182910/2022|EPI_ISL_10666994|2022−01−17
Japan/PG−182902/2022|EPI_ISL_10666986|2022−01−14
Japan/PG−182896/2022|EPI_ISL_10666980|2022−01−13
Japan/PG−180750/2022|EPI_ISL_10666965|2022−01−13
Japan/PG−180742/2022|EPI_ISL_10666958|2022−01−12
Japan/PG−182943/2022|EPI_ISL_10667239|2022−01−14
Japan/YCH1096/2022|EPI_ISL_10115930|2022−01−22
Japan/YCH1087/2022|EPI_ISL_10115921|2022−02−14
Japan/YCH1069/2022|EPI_ISL_10067786|2022−01−20
Japan/YCH1057/2022|EPI_ISL_9847997|2022−02−07
Japan/TKYkbm9578/2022|EPI_ISL_11172850|2022−02−10
Japan/YCH1041/2022|EPI_ISL_9640044|2022−01−17
Japan/PG−201320/2022|EPI_ISL_11712099|2022−02−18
Japan/YCH1228_1/2022|EPI_ISL_11050901|2022−02−18
Japan/PG−192234/2022|EPI_ISL_10667339|2022−02−04
Japan/YCH1085/2022|EPI_ISL_10115919|2022−02−12
Japan/YCH0937/2022|EPI_ISL_9166485|2022−01−22
0
10
20
30
40
50
Australia/VIC63916/2022|EPI_ISL_13709076|2022−06−03
Australia/VIC64068/2022|EPI_ISL_13709241|2022−06−04
C-to-T: 11, G-to-A: 9, T-to-C: 3
Australia/VIC64069/2022|EPI_ISL_13709242|2022−06−04
Australia/VIC64105/2022|EPI_ISL_13709277|2022−06−04
Australia/VIC64070/2022|EPI_ISL_13709243|2022−06−04
Australia/VIC60339/2022|EPI_ISL_13274389|2022−05−11
Australia/VIC59446/2022|EPI_ISL_13274388|2022−05−11
Australia/VIC59444/2022|EPI_ISL_13274386|2022−05−11
Australia/VIC60999/2022|EPI_ISL_13457287|2022−05−24
Australia/VIC58815/2022|EPI_ISL_12979587|2022−05−08
Australia/VIC63337/2022|EPI_ISL_13458410|2022−06−03
Australia/VIC61778/2022|EPI_ISL_13276482|2022−05−28
0
5
10
15
20
25
Sweden/1592741163/2022|EPI_ISL_12916821|2022−05−09
Sweden/1592739463/2022|EPI_ISL_12916827|2022−05−06
Sweden/1592740963/2022|EPI_ISL_12916846|2022−05−05
C-to-T: 4, G-to-A: 3, T-to-C: 2, A-to-G: 1, C-to-A: 1
Sweden/1592738063/2022|EPI_ISL_12916848|2022−05−06
Sweden/1596751663/2022|EPI_ISL_12916820|2022−05−02
Sweden/1592331163/2022|EPI_ISL_12265678|2022−03−31
0
5
10
15

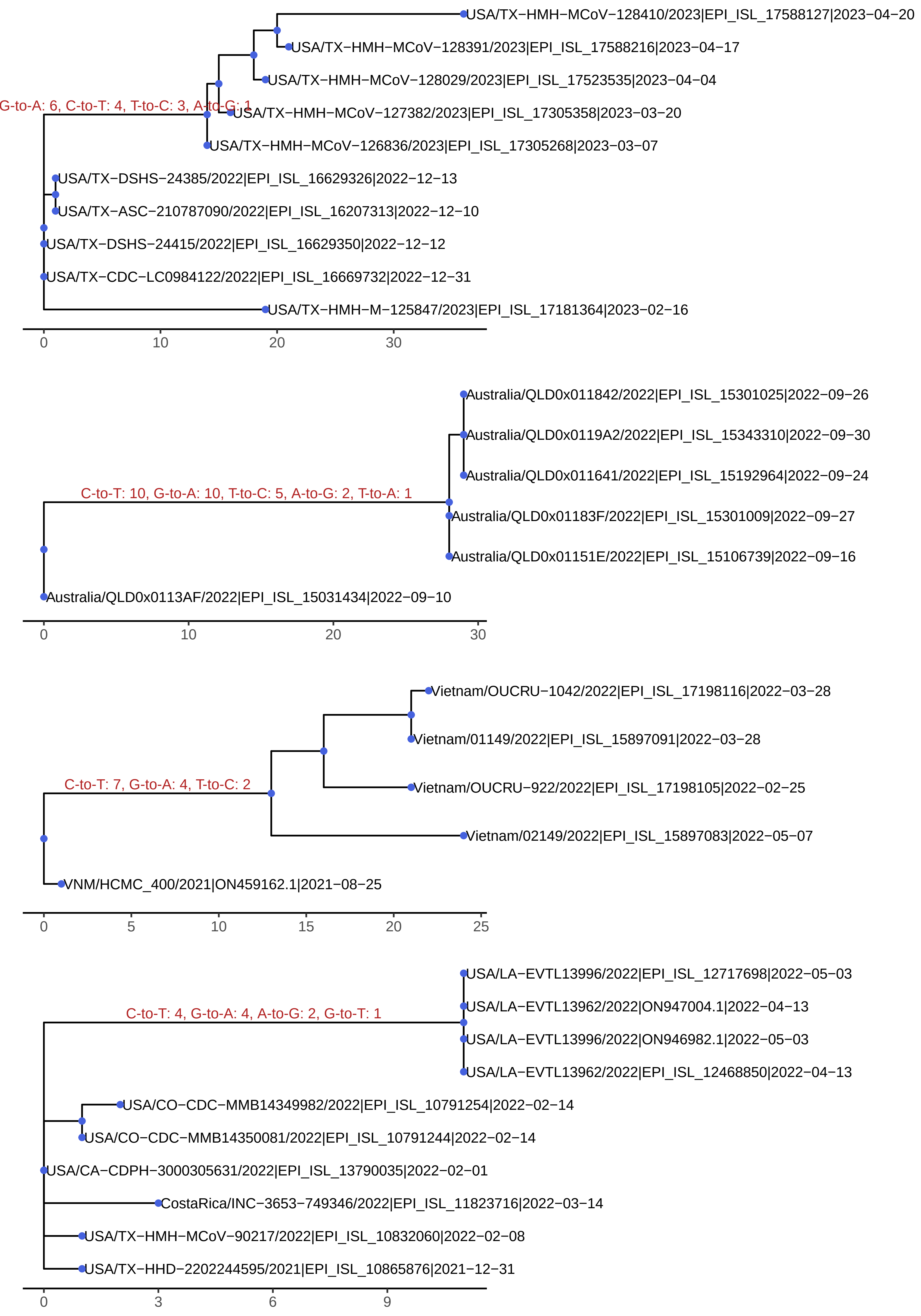
USA/TX−HMH−MCoV−128410/2023|EPI_ISL_17588127|2023−04−20
USA/TX−HMH−MCoV−128391/2023|EPI_ISL_17588216|2023−04−17
USA/TX−HMH−MCoV−128029/2023|EPI_ISL_17523535|2023−04−04
G-to-A: 6, C-to-T: 4, T-to-C: 3, A-to-G: 1
USA/TX−HMH−MCoV−127382/2023|EPI_ISL_17305358|2023−03−20
USA/TX−HMH−MCoV−126836/2023|EPI_ISL_17305268|2023−03−07
USA/TX−DSHS−24385/2022|EPI_ISL_16629326|2022−12−13
USA/TX−ASC−210787090/2022|EPI_ISL_16207313|2022−12−10
USA/TX−DSHS−24415/2022|EPI_ISL_16629350|2022−12−12
USA/TX−CDC−LC0984122/2022|EPI_ISL_16669732|2022−12−31
USA/TX−HMH−M−125847/2023|EPI_ISL_17181364|2023−02−16
0
10
20
30
Australia/QLD0x011842/2022|EPI_ISL_15301025|2022−09−26
Australia/QLD0x0119A2/2022|EPI_ISL_15343310|2022−09−30
Australia/QLD0x011641/2022|EPI_ISL_15192964|2022−09−24
C-to-T: 10, G-to-A: 10, T-to-C: 5, A-to-G: 2, T-to-A: 1
Australia/QLD0x01183F/2022|EPI_ISL_15301009|2022−09−27
Australia/QLD0x01151E/2022|EPI_ISL_15106739|2022−09−16
Australia/QLD0x0113AF/2022|EPI_ISL_15031434|2022−09−10
0
10
20
30
Vietnam/OUCRU−1042/2022|EPI_ISL_17198116|2022−03−28
Vietnam/01149/2022|EPI_ISL_15897091|2022−03−28
C-to-T: 7, G-to-A: 4, T-to-C: 2
Vietnam/OUCRU−922/2022|EPI_ISL_17198105|2022−02−25
Vietnam/02149/2022|EPI_ISL_15897083|2022−05−07
VNM/HCMC_400/2021|ON459162.1|2021−08−25
0
5
10
15
20
25
USA/LA−EVTL13996/2022|EPI_ISL_12717698|2022−05−03
C-to-T: 4, G-to-A: 4, A-to-G: 2, G-to-T: 1
USA/LA−EVTL13962/2022|ON947004.1|2022−04−13
USA/LA−EVTL13996/2022|ON946982.1|2022−05−03
USA/LA−EVTL13962/2022|EPI_ISL_12468850|2022−04−13
USA/CO−CDC−MMB14349982/2022|EPI_ISL_10791254|2022−02−14
USA/CO−CDC−MMB14350081/2022|EPI_ISL_10791244|2022−02−14
USA/CA−CDPH−3000305631/2022|EPI_ISL_13790035|2022−02−01
CostaRica/INC−3653−749346/2022|EPI_ISL_11823716|2022−03−14
USA/TX−HMH−MCoV−90217/2022|EPI_ISL_10832060|2022−02−08
USA/TX−HHD−2202244595/2021|EPI_ISL_10865876|2021−12−31
0
3
6
9

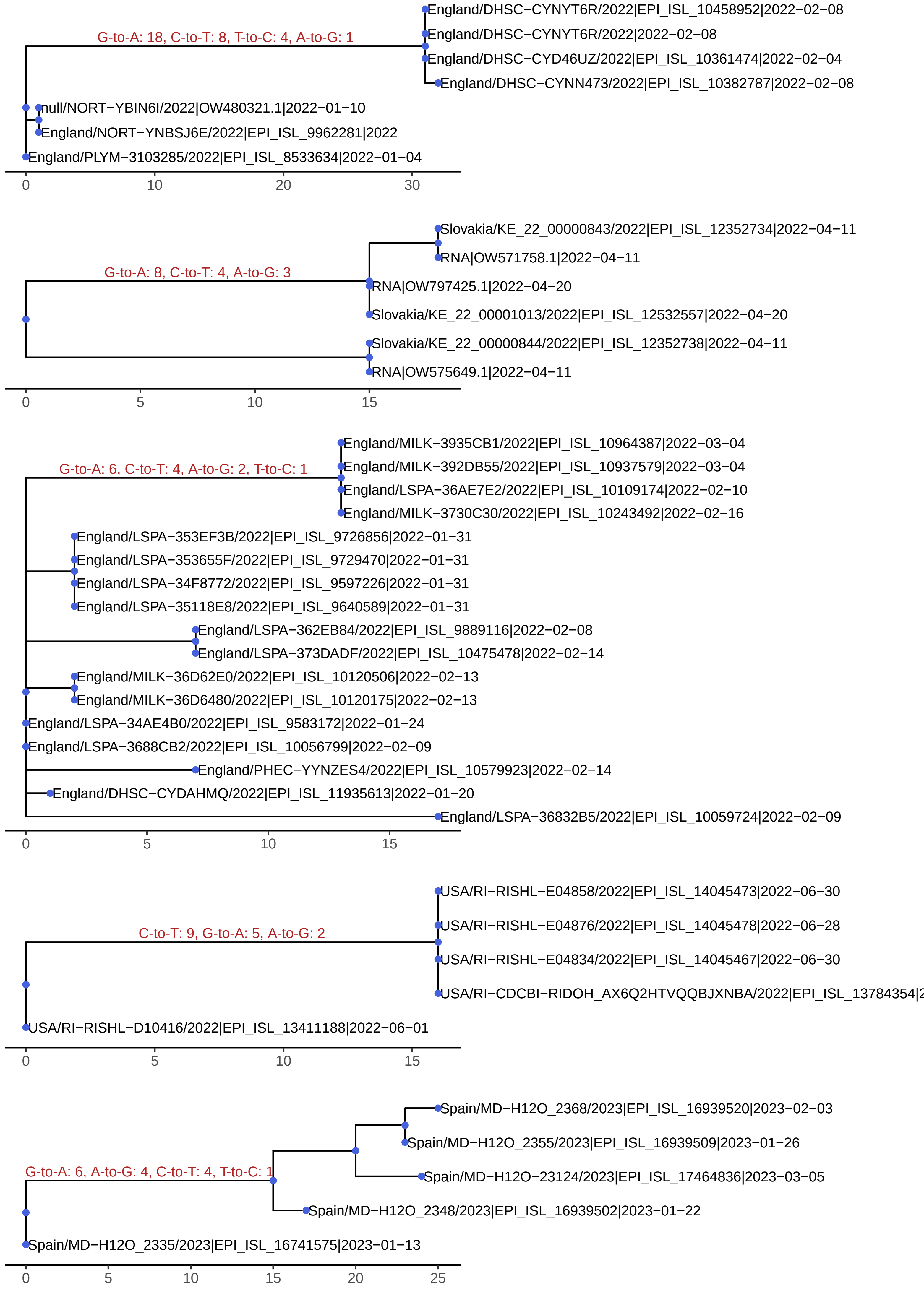
G-to-A: 18, C-to-T: 8, T-to-C: 4, A-to-G: 1
England/DHSC−CYNYT6R/2022|EPI_ISL_10458952|2022−02−08
England/DHSC−CYNYT6R/2022|2022−02−08
England/DHSC−CYD46UZ/2022|EPI_ISL_10361474|2022−02−04
England/DHSC−CYNN473/2022|EPI_ISL_10382787|2022−02−08
null/NORT−YBIN6I/2022|OW480321.1|2022−01−10
England/NORT−YNBSJ6E/2022|EPI_ISL_9962281|2022
England/PLYM−3103285/2022|EPI_ISL_8533634|2022−01−04
0
10
20
30
G-to-A: 8, C-to-T: 4, A-to-G: 3
Slovakia/KE_22_00000843/2022|EPI_ISL_12352734|2022−04−11
RNA|OW571758.1|2022−04−11
RNA|OW797425.1|2022−04−20
Slovakia/KE_22_00001013/2022|EPI_ISL_12532557|2022−04−20
Slovakia/KE_22_00000844/2022|EPI_ISL_12352738|2022−04−11
RNA|OW575649.1|2022−04−11
0
5
10
15
G-to-A: 6, C-to-T: 4, A-to-G: 2, T-to-C: 1
England/MILK−3935CB1/2022|EPI_ISL_10964387|2022−03−04
England/MILK−392DB55/2022|EPI_ISL_10937579|2022−03−04
England/LSPA−36AE7E2/2022|EPI_ISL_10109174|2022−02−10
England/MILK−3730C30/2022|EPI_ISL_10243492|2022−02−16
England/LSPA−353EF3B/2022|EPI_ISL_9726856|2022−01−31
England/LSPA−353655F/2022|EPI_ISL_9729470|2022−01−31
England/LSPA−34F8772/2022|EPI_ISL_9597226|2022−01−31
England/LSPA−35118E8/2022|EPI_ISL_9640589|2022−01−31
England/LSPA−362EB84/2022|EPI_ISL_9889116|2022−02−08
England/LSPA−373DADF/2022|EPI_ISL_10475478|2022−02−14
England/MILK−36D62E0/2022|EPI_ISL_10120506|2022−02−13
England/MILK−36D6480/2022|EPI_ISL_10120175|2022−02−13
England/LSPA−34AE4B0/2022|EPI_ISL_9583172|2022−01−24
England/LSPA−3688CB2/2022|EPI_ISL_10056799|2022−02−09
England/PHEC−YYNZES4/2022|EPI_ISL_10579923|2022−02−14
England/DHSC−CYDAHMQ/2022|EPI_ISL_11935613|2022−01−20
England/LSPA−36832B5/2022|EPI_ISL_10059724|2022−02−09
0
5
10
15
C-to-T: 9, G-to-A: 5, A-to-G: 2
USA/RI−RISHL−E04858/2022|EPI_ISL_14045473|2022−06−30
USA/RI−RISHL−E04876/2022|EPI_ISL_14045478|2022−06−28
USA/RI−RISHL−E04834/2022|EPI_ISL_14045467|2022−06−30
USA/RI−CDCBI−RIDOH_AX6Q2HTVQQBJXNBA/2022|EPI_ISL_13784354|2
USA/RI−RISHL−D10416/2022|EPI_ISL_13411188|2022−06−01
0
5
10
15
G-to-A: 6, A-to-G: 4, C-to-T: 4, T-to-C: 1
Spain/MD−H12O_2368/2023|EPI_ISL_16939520|2023−02−03
Spain/MD−H12O_2355/2023|EPI_ISL_16939509|2023−01−26
Spain/MD−H12O−23124/2023|EPI_ISL_17464836|2023−03−05
Spain/MD−H12O_2348/2023|EPI_ISL_16939502|2023−01−22
Spain/MD−H12O_2335/2023|EPI_ISL_16741575|2023−01−13
0
5
10
15
20
25

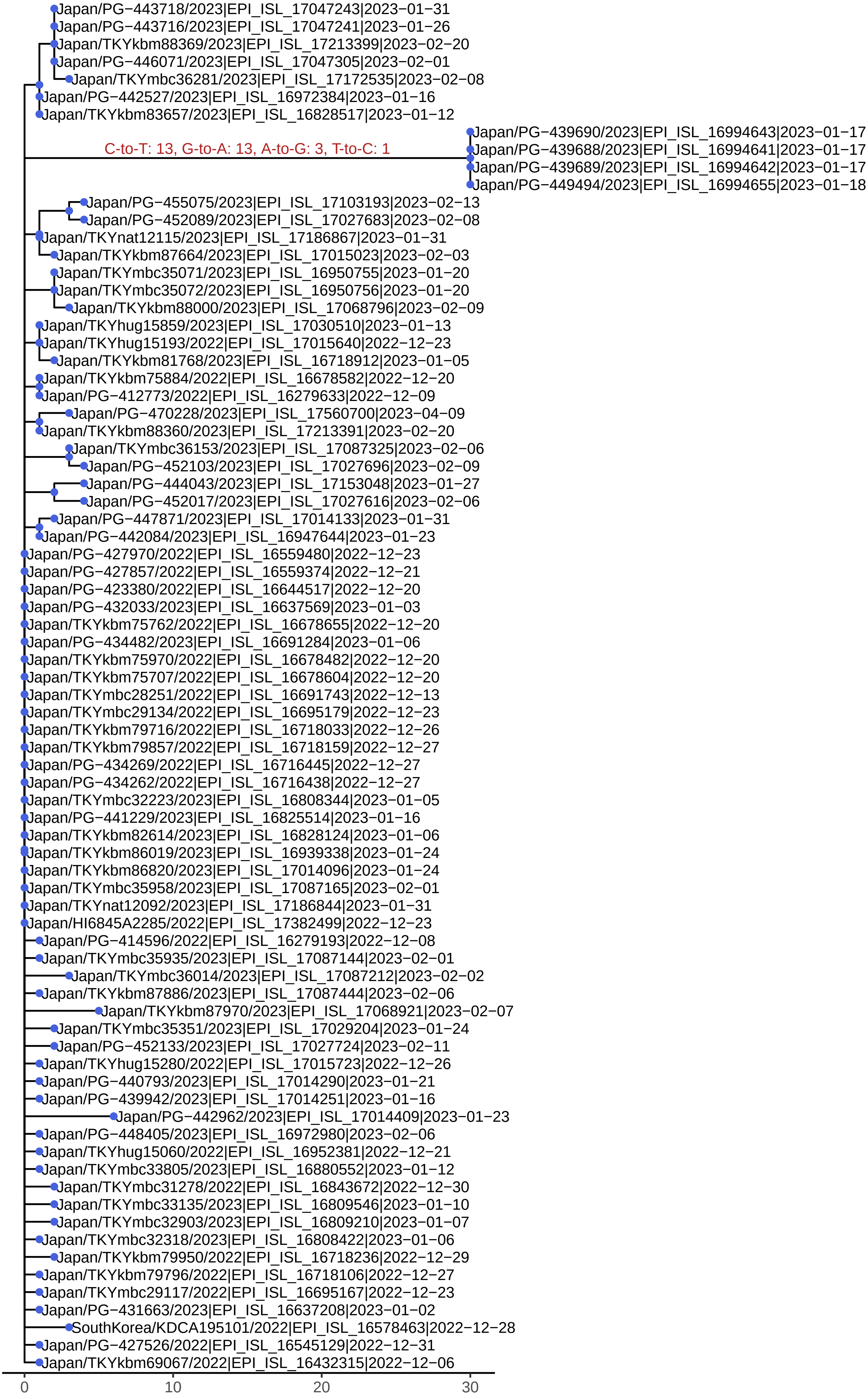

Japan/PG−443718/2023|EPI_ISL_17047243|2023−01−31
Japan/PG−443716/2023|EPI_ISL_17047241|2023−01−26
Japan/TKYkbm88369/2023|EPI_ISL_17213399|2023−02−20
Japan/PG−446071/2023|EPI_ISL_17047305|2023−02−01
Japan/TKYmbc36281/2023|EPI_ISL_17172535|2023−02−08
Japan/PG−442527/2023|EPI_ISL_16972384|2023−01−16
Japan/TKYkbm83657/2023|EPI_ISL_16828517|2023−01−12
Japan/PG−439690/2023|EPI_ISL_16994643|2023−01−17
C-to-T: 13, G-to-A: 13, A-to-G: 3, T-to-C: 1
Japan/PG−439688/2023|EPI_ISL_16994641|2023−01−17
Japan/PG−439689/2023|EPI_ISL_16994642|2023−01−17
Japan/PG−449494/2023|EPI_ISL_16994655|2023−01−18
Japan/PG−455075/2023|EPI_ISL_17103193|2023−02−13
Japan/PG−452089/2023|EPI_ISL_17027683|2023−02−08
Japan/TKYnat12115/2023|EPI_ISL_17186867|2023−01−31
Japan/TKYkbm87664/2023|EPI_ISL_17015023|2023−02−03
Japan/TKYmbc35071/2023|EPI_ISL_16950755|2023−01−20
Japan/TKYmbc35072/2023|EPI_ISL_16950756|2023−01−20
Japan/TKYkbm88000/2023|EPI_ISL_17068796|2023−02−09
Japan/TKYhug15859/2023|EPI_ISL_17030510|2023−01−13
Japan/TKYhug15193/2022|EPI_ISL_17015640|2022−12−23
Japan/TKYkbm81768/2023|EPI_ISL_16718912|2023−01−05
Japan/TKYkbm75884/2022|EPI_ISL_16678582|2022−12−20
Japan/PG−412773/2022|EPI_ISL_16279633|2022−12−09
Japan/PG−470228/2023|EPI_ISL_17560700|2023−04−09
Japan/TKYkbm88360/2023|EPI_ISL_17213391|2023−02−20
Japan/TKYmbc36153/2023|EPI_ISL_17087325|2023−02−06
Japan/PG−452103/2023|EPI_ISL_17027696|2023−02−09
Japan/PG−444043/2023|EPI_ISL_17153048|2023−01−27
Japan/PG−452017/2023|EPI_ISL_17027616|2023−02−06
Japan/PG−447871/2023|EPI_ISL_17014133|2023−01−31
Japan/PG−442084/2023|EPI_ISL_16947644|2023−01−23
Japan/PG−427970/2022|EPI_ISL_16559480|2022−12−23
Japan/PG−427857/2022|EPI_ISL_16559374|2022−12−21
Japan/PG−423380/2022|EPI_ISL_16644517|2022−12−20
Japan/PG−432033/2023|EPI_ISL_16637569|2023−01−03
Japan/TKYkbm75762/2022|EPI_ISL_16678655|2022−12−20
Japan/PG−434482/2023|EPI_ISL_16691284|2023−01−06
Japan/TKYkbm75970/2022|EPI_ISL_16678482|2022−12−20
Japan/TKYkbm75707/2022|EPI_ISL_16678604|2022−12−20
Japan/TKYmbc28251/2022|EPI_ISL_16691743|2022−12−13
Japan/TKYmbc29134/2022|EPI_ISL_16695179|2022−12−23
Japan/TKYkbm79716/2022|EPI_ISL_16718033|2022−12−26
Japan/TKYkbm79857/2022|EPI_ISL_16718159|2022−12−27
Japan/PG−434269/2022|EPI_ISL_16716445|2022−12−27
Japan/PG−434262/2022|EPI_ISL_16716438|2022−12−27
Japan/TKYmbc32223/2023|EPI_ISL_16808344|2023−01−05
Japan/PG−441229/2023|EPI_ISL_16825514|2023−01−16
Japan/TKYkbm82614/2023|EPI_ISL_16828124|2023−01−06
Japan/TKYkbm86019/2023|EPI_ISL_16939338|2023−01−24
Japan/TKYkbm86820/2023|EPI_ISL_17014096|2023−01−24
Japan/TKYmbc35958/2023|EPI_ISL_17087165|2023−02−01
Japan/TKYnat12092/2023|EPI_ISL_17186844|2023−01−31
Japan/HI6845A2285/2022|EPI_ISL_17382499|2022−12−23
Japan/PG−414596/2022|EPI_ISL_16279193|2022−12−08
Japan/TKYmbc35935/2023|EPI_ISL_17087144|2023−02−01
Japan/TKYmbc36014/2023|EPI_ISL_17087212|2023−02−02
Japan/TKYkbm87886/2023|EPI_ISL_17087444|2023−02−06
Japan/TKYkbm87970/2023|EPI_ISL_17068921|2023−02−07
Japan/TKYmbc35351/2023|EPI_ISL_17029204|2023−01−24
Japan/PG−452133/2023|EPI_ISL_17027724|2023−02−11
Japan/TKYhug15280/2022|EPI_ISL_17015723|2022−12−26
Japan/PG−440793/2023|EPI_ISL_17014290|2023−01−21
Japan/PG−439942/2023|EPI_ISL_17014251|2023−01−16
Japan/PG−442962/2023|EPI_ISL_17014409|2023−01−23
Japan/PG−448405/2023|EPI_ISL_16972980|2023−02−06
Japan/TKYhug15060/2022|EPI_ISL_16952381|2022−12−21
Japan/TKYmbc33805/2023|EPI_ISL_16880552|2023−01−12
Japan/TKYmbc31278/2022|EPI_ISL_16843672|2022−12−30
Japan/TKYmbc33135/2023|EPI_ISL_16809546|2023−01−10
Japan/TKYmbc32903/2023|EPI_ISL_16809210|2023−01−07
Japan/TKYmbc32318/2023|EPI_ISL_16808422|2023−01−06
Japan/TKYkbm79950/2022|EPI_ISL_16718236|2022−12−29
Japan/TKYkbm79796/2022|EPI_ISL_16718106|2022−12−27
Japan/TKYmbc29117/2022|EPI_ISL_16695167|2022−12−23
Japan/PG−431663/2023|EPI_ISL_16637208|2023−01−02
SouthKorea/KDCA195101/2022|EPI_ISL_16578463|2022−12−28
Japan/PG−427526/2022|EPI_ISL_16545129|2022−12−31
Japan/TKYkbm69067/2022|EPI_ISL_16432315|2022−12−06
0
10
20
30

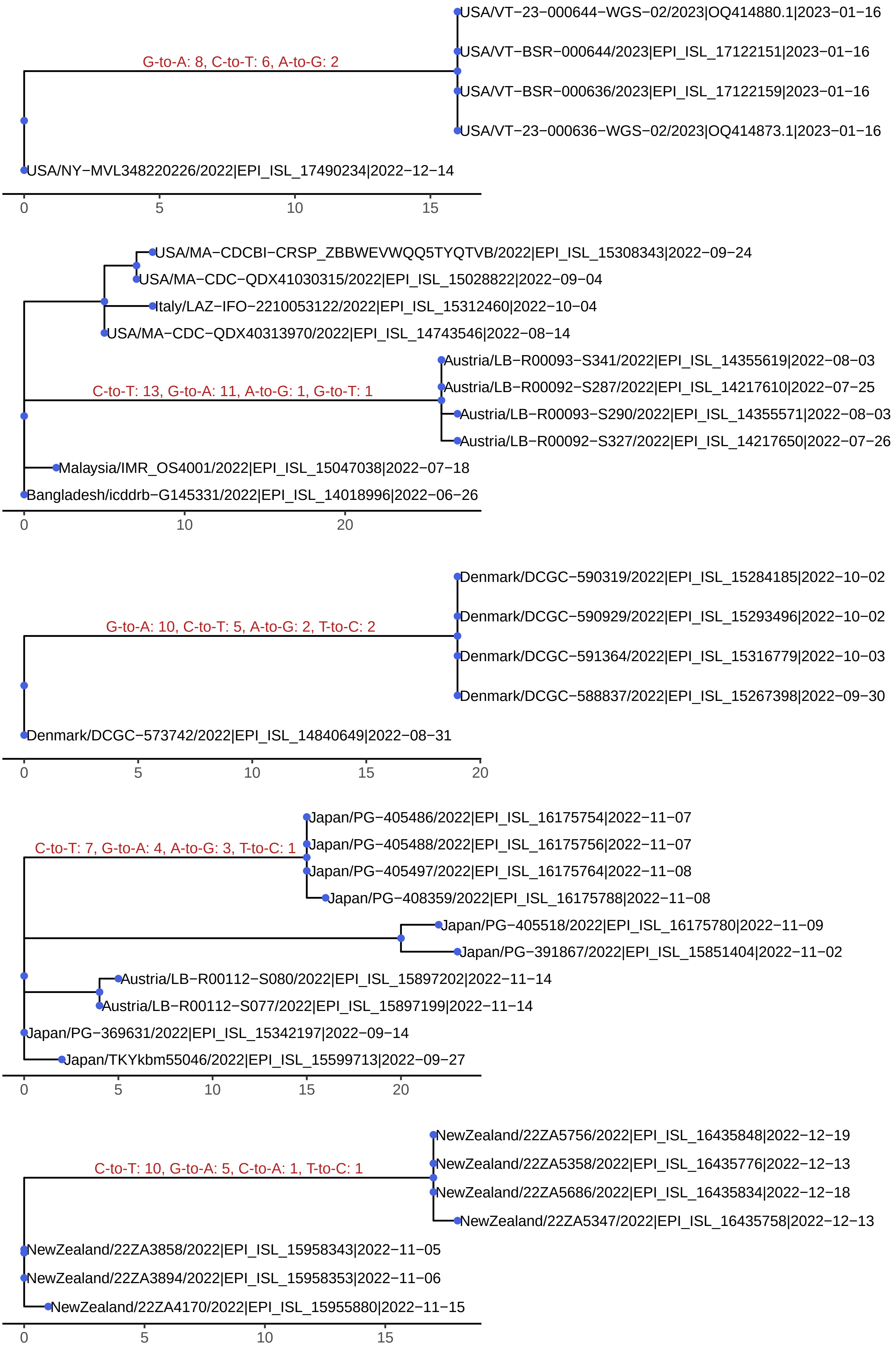

G-to-A: 8, C-to-T: 6, A-to-G: 2
USA/VT−23−000644−WGS−02/2023|OQ414880.1|2023−01−16
USA/VT−BSR−000644/2023|EPI_ISL_17122151|2023−01−16
USA/VT−BSR−000636/2023|EPI_ISL_17122159|2023−01−16
USA/VT−23−000636−WGS−02/2023|OQ414873.1|2023−01−16
USA/NY−MVL348220226/2022|EPI_ISL_17490234|2022−12−14
0 5 10 15
USA/MA−CDCBI−CRSP_ZBBWEVWQQ5TYQTVB/2022|EPI_ISL_15308343|2022−09−24
USA/MA−CDC−QDX41030315/2022|EPI_ISL_15028822|2022−09−04
Italy/LAZ−IFO−2210053122/2022|EPI_ISL_15312460|2022−10−04
USA/MA−CDC−QDX40313970/2022|EPI_ISL_14743546|2022−08−14
C-to-T: 13, G-to-A: 11, A-to-G: 1, G-to-T: 1
Austria/LB−R00093−S341/2022|EPI_ISL_14355619|2022−08−03
Austria/LB−R00092−S287/2022|EPI_ISL_14217610|2022−07−25
Austria/LB−R00093−S290/2022|EPI_ISL_14355571|2022−08−03
Austria/LB−R00092−S327/2022|EPI_ISL_14217650|2022−07−26
Malaysia/IMR_OS4001/2022|EPI_ISL_15047038|2022−07−18
Bangladesh/icddrb−G145331/2022|EPI_ISL_14018996|2022−06−26
0 10 20
G-to-A: 10, C-to-T: 5, A-to-G: 2, T-to-C: 2
Denmark/DCGC−590319/2022|EPI_ISL_15284185|2022−10−02
Denmark/DCGC−590929/2022|EPI_ISL_15293496|2022−10−02
Denmark/DCGC−591364/2022|EPI_ISL_15316779|2022−10−03
Denmark/DCGC−588837/2022|EPI_ISL_15267398|2022−09−30
Denmark/DCGC−573742/2022|EPI_ISL_14840649|2022−08−31
0 5 10 15 20
C-to-T: 7, G-to-A: 4, A-to-G: 3, T-to-C: 1
Japan/PG−405486/2022|EPI_ISL_16175754|2022−11−07
Japan/PG−405488/2022|EPI_ISL_16175756|2022−11−07
Japan/PG−405497/2022|EPI_ISL_16175764|2022−11−08
Japan/PG−408359/2022|EPI_ISL_16175788|2022−11−08
Japan/PG−405518/2022|EPI_ISL_16175780|2022−11−09
Japan/PG−391867/2022|EPI_ISL_15851404|2022−11−02
Austria/LB−R00112−S080/2022|EPI_ISL_15897202|2022−11−14
Austria/LB−R00112−S077/2022|EPI_ISL_15897199|2022−11−14
Japan/PG−369631/2022|EPI_ISL_15342197|2022−09−14
Japan/TKYkbm55046/2022|EPI_ISL_15599713|2022−09−27
0 5 10 15 20
C-to-T: 10, G-to-A: 5, C-to-A: 1, T-to-C: 1
NewZealand/22ZA5756/2022|EPI_ISL_16435848|2022−12−19
NewZealand/22ZA5358/2022|EPI_ISL_16435776|2022−12−13
NewZealand/22ZA5686/2022|EPI_ISL_16435834|2022−12−18
NewZealand/22ZA5347/2022|EPI_ISL_16435758|2022−12−13
NewZealand/22ZA3858/2022|EPI_ISL_15958343|2022−11−05
NewZealand/22ZA3894/2022|EPI_ISL_15958353|2022−11−06
NewZealand/22ZA4170/2022|EPI_ISL_15955880|2022−11−15
0 5 10 15

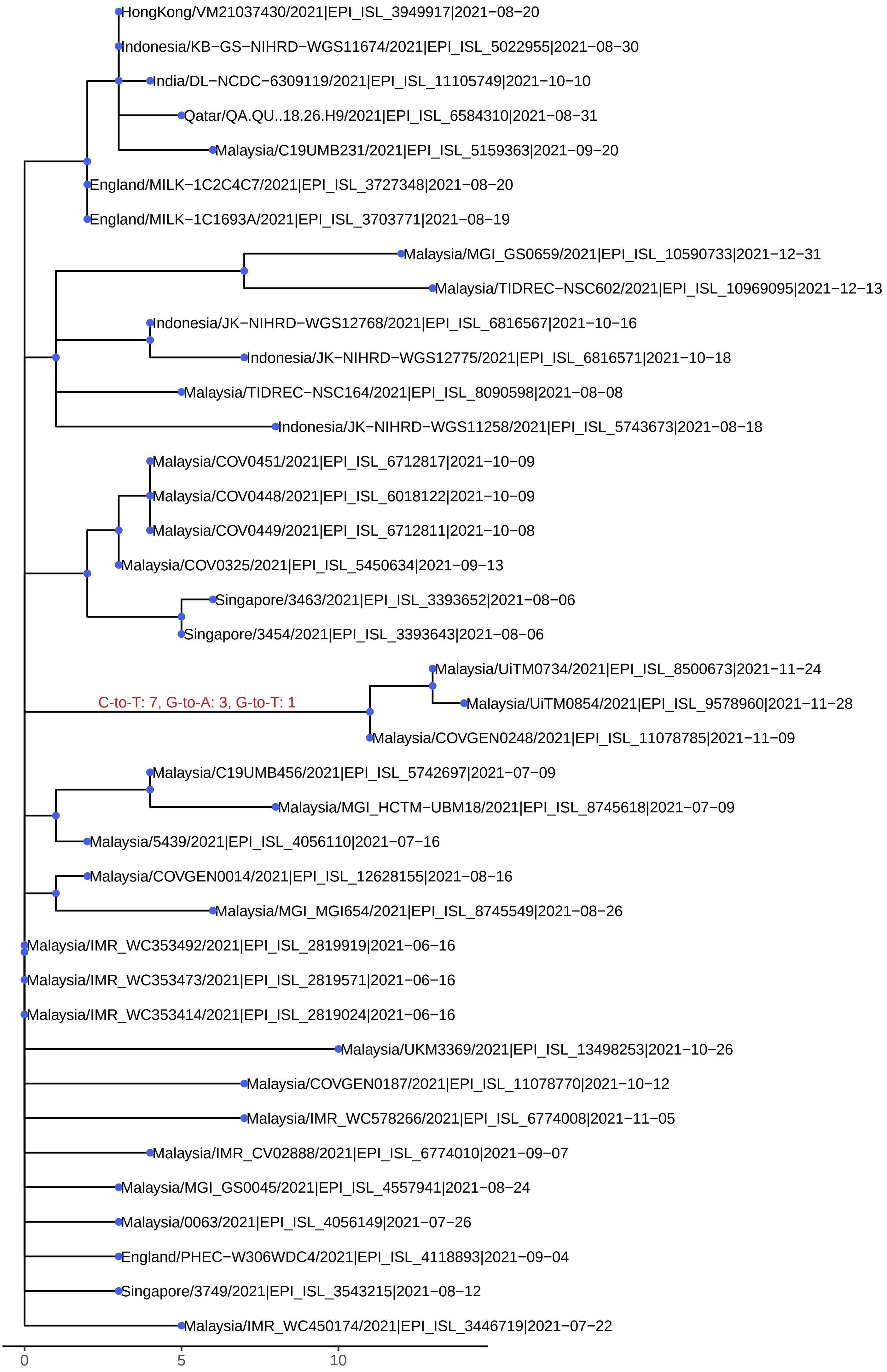

HongKong/VM21037430/2021|EPI_ISL_3949917|2021−08−20
Indonesia/KB−GS−NIHRD−WGS11674/2021|EPI_ISL_5022955|2021−08−30
India/DL−NCDC−6309119/2021|EPI_ISL_11105749|2021−10−10
Qatar/QA.QU..18.26.H9/2021|EPI_ISL_6584310|2021−08−31
Malaysia/C19UMB231/2021|EPI_ISL_5159363|2021−09−20
England/MILK−1C2C4C7/2021|EPI_ISL_3727348|2021−08−20
England/MILK−1C1693A/2021|EPI_ISL_3703771|2021−08−19
Malaysia/MGI_GS0659/2021|EPI_ISL_10590733|2021−12−31
Malaysia/TIDREC−NSC602/2021|EPI_ISL_10969095|2021−12−13
Indonesia/JK−NIHRD−WGS12768/2021|EPI_ISL_6816567|2021−10−16
Indonesia/JK−NIHRD−WGS12775/2021|EPI_ISL_6816571|2021−10−18
Malaysia/TIDREC−NSC164/2021|EPI_ISL_8090598|2021−08−08
Indonesia/JK−NIHRD−WGS11258/2021|EPI_ISL_5743673|2021−08−18
Malaysia/COV0451/2021|EPI_ISL_6712817|2021−10−09
Malaysia/COV0448/2021|EPI_ISL_6018122|2021−10−09
Malaysia/COV0449/2021|EPI_ISL_6712811|2021−10−08
Malaysia/COV0325/2021|EPI_ISL_5450634|2021−09−13
Singapore/3463/2021|EPI_ISL_3393652|2021−08−06
Singapore/3454/2021|EPI_ISL_3393643|2021−08−06
Malaysia/UiTM0734/2021|EPI_ISL_8500673|2021−11−24
C-to-T: 7, G-to-A: 3, G-to-T: 1
Malaysia/UiTM0854/2021|EPI_ISL_9578960|2021−11−28
Malaysia/COVGEN0248/2021|EPI_ISL_11078785|2021−11−09
Malaysia/C19UMB456/2021|EPI_ISL_5742697|2021−07−09
Malaysia/MGI_HCTM−UBM18/2021|EPI_ISL_8745618|2021−07−09
Malaysia/5439/2021|EPI_ISL_4056110|2021−07−16
Malaysia/COVGEN0014/2021|EPI_ISL_12628155|2021−08−16
Malaysia/MGI_MGI654/2021|EPI_ISL_8745549|2021−08−26
Malaysia/IMR_WC353492/2021|EPI_ISL_2819919|2021−06−16
Malaysia/IMR_WC353473/2021|EPI_ISL_2819571|2021−06−16
Malaysia/IMR_WC353414/2021|EPI_ISL_2819024|2021−06−16
Malaysia/UKM3369/2021|EPI_ISL_13498253|2021−10−26
Malaysia/COVGEN0187/2021|EPI_ISL_11078770|2021−10−12
Malaysia/IMR_WC578266/2021|EPI_ISL_6774008|2021−11−05
Malaysia/IMR_CV02888/2021|EPI_ISL_6774010|2021−09−07
Malaysia/MGI_GS0045/2021|EPI_ISL_4557941|2021−08−24
Malaysia/0063/2021|EPI_ISL_4056149|2021−07−26
England/PHEC−W306WDC4/2021|EPI_ISL_4118893|2021−09−04
Singapore/3749/2021|EPI_ISL_3543215|2021−08−12
Malaysia/IMR_WC450174/2021|EPI_ISL_3446719|2021−07−22
0
5
10

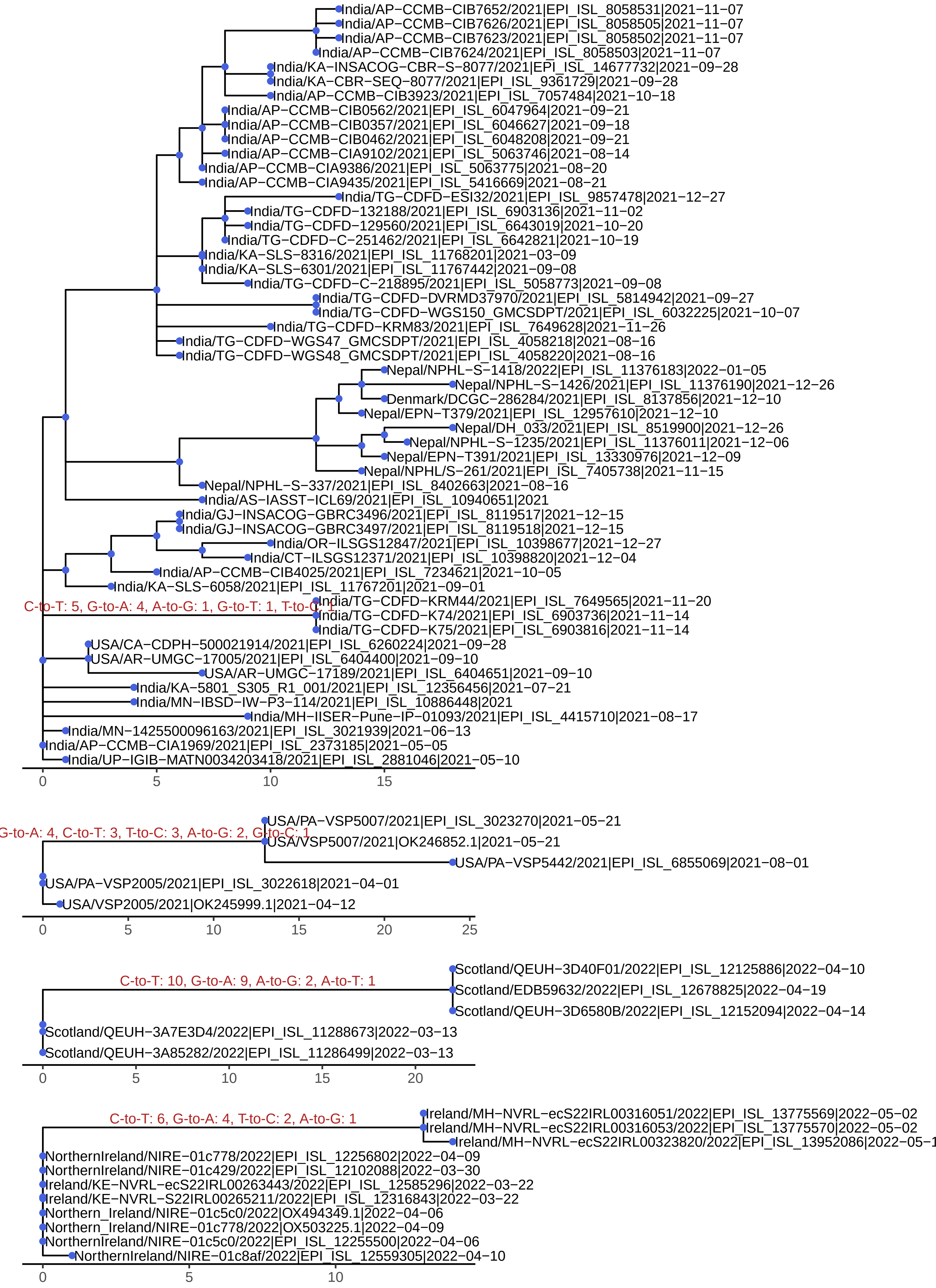

India/AP−CCMB−CIB7652/2021|EPI_ISL_8058531|2021−11−07
India/AP−CCMB−CIB7626/2021|EPI_ISL_8058505|2021−11−07
India/AP−CCMB−CIB7623/2021|EPI_ISL_8058502|2021−11−07
India/AP−CCMB−CIB7624/2021|EPI_ISL_8058503|2021−11−07
India/KA−INSACOG−CBR−S−8077/2021|EPI_ISL_14677732|2021−09−28
India/KA−CBR−SEQ−8077/2021|EPI_ISL_9361729|2021−09−28
India/AP−CCMB−CIB3923/2021|EPI_ISL_7057484|2021−10−18
India/AP−CCMB−CIB0562/2021|EPI_ISL_6047964|2021−09−21
India/AP−CCMB−CIB0357/2021|EPI_ISL_6046627|2021−09−18
India/AP−CCMB−CIB0462/2021|EPI_ISL_6048208|2021−09−21
India/AP−CCMB−CIA9102/2021|EPI_ISL_5063746|2021−08−14
India/AP−CCMB−CIA9386/2021|EPI_ISL_5063775|2021−08−20
India/AP−CCMB−CIA9435/2021|EPI_ISL_5416669|2021−08−21
India/TG−CDFD−ESI32/2021|EPI_ISL_9857478|2021−12−27
India/TG−CDFD−132188/2021|EPI_ISL_6903136|2021−11−02
India/TG−CDFD−129560/2021|EPI_ISL_6643019|2021−10−20
India/TG−CDFD−C−251462/2021|EPI_ISL_6642821|2021−10−19
India/KA−SLS−8316/2021|EPI_ISL_11768201|2021−03−09
India/KA−SLS−6301/2021|EPI_ISL_11767442|2021−09−08
India/TG−CDFD−C−218895/2021|EPI_ISL_5058773|2021−09−08
India/TG−CDFD−DVRMD37970/2021|EPI_ISL_5814942|2021−09−27
India/TG−CDFD−WGS150_GMCSDPT/2021|EPI_ISL_6032225|2021−10−07
India/TG−CDFD−KRM83/2021|EPI_ISL_7649628|2021−11−26
India/TG−CDFD−WGS47_GMCSDPT/2021|EPI_ISL_4058218|2021−08−16
India/TG−CDFD−WGS48_GMCSDPT/2021|EPI_ISL_4058220|2021−08−16
Nepal/NPHL−S−1418/2022|EPI_ISL_11376183|2022−01−05
Nepal/NPHL−S−1426/2021|EPI_ISL_11376190|2021−12−26
Denmark/DCGC−286284/2021|EPI_ISL_8137856|2021−12−10
Nepal/EPN−T379/2021|EPI_ISL_12957610|2021−12−10
Nepal/DH_033/2021|EPI_ISL_8519900|2021−12−26
Nepal/NPHL−S−1235/2021|EPI_ISL_11376011|2021−12−06
Nepal/EPN−T391/2021|EPI_ISL_13330976|2021−12−09
Nepal/NPHL/S−261/2021|EPI_ISL_7405738|2021−11−15
Nepal/NPHL−S−337/2021|EPI_ISL_8402663|2021−08−16
India/AS−IASST−ICL69/2021|EPI_ISL_10940651|2021
India/GJ−INSACOG−GBRC3496/2021|EPI_ISL_8119517|2021−12−15
India/GJ−INSACOG−GBRC3497/2021|EPI_ISL_8119518|2021−12−15
India/OR−ILSGS12847/2021|EPI_ISL_10398677|2021−12−27
India/CT−ILSGS12371/2021|EPI_ISL_10398820|2021−12−04
India/AP−CCMB−CIB4025/2021|EPI_ISL_7234621|2021−10−05
India/KA−SLS−6058/2021|EPI_ISL_11767201|2021−09−01
C-to-T: 5, G-to-A: 4, A-to-G: 1, G-to-T: 1, T-to-C: 1
India/TG−CDFD−KRM44/2021|EPI_ISL_7649565|2021−11−20
India/TG−CDFD−K74/2021|EPI_ISL_6903736|2021−11−14
India/TG−CDFD−K75/2021|EPI_ISL_6903816|2021−11−14
USA/CA−CDPH−500021914/2021|EPI_ISL_6260224|2021−09−28
USA/AR−UMGC−17005/2021|EPI_ISL_6404400|2021−09−10
USA/AR−UMGC−17189/2021|EPI_ISL_6404651|2021−09−10
India/KA−5801_S305_R1_001/2021|EPI_ISL_12356456|2021−07−21
India/MN−IBSD−IW−P3−114/2021|EPI_ISL_10886448|2021
India/MH−IISER−Pune−IP−01093/2021|EPI_ISL_4415710|2021−08−17
India/MN−1425500096163/2021|EPI_ISL_3021939|2021−06−13
India/AP−CCMB−CIA1969/2021|EPI_ISL_2373185|2021−05−05
India/UP−IGIB−MATN0034203418/2021|EPI_ISL_2881046|2021−05−10
0
5
10
15
G-to-A: 4, C-to-T: 3, T-to-C: 3, A-to-G: 2, G-to-C: 1
USA/PA−VSP5007/2021|EPI_ISL_3023270|2021−05−21
USA/VSP5007/2021|OK246852.1|2021−05−21
USA/PA−VSP5442/2021|EPI_ISL_6855069|2021−08−01
USA/PA−VSP2005/2021|EPI_ISL_3022618|2021−04−01
USA/VSP2005/2021|OK245999.1|2021−04−12
0
5
10
15
20
25
C-to-T: 10, G-to-A: 9, A-to-G: 2, A-to-T: 1
Scotland/QEUH−3D40F01/2022|EPI_ISL_12125886|2022−04−10
Scotland/EDB59632/2022|EPI_ISL_12678825|2022−04−19
Scotland/QEUH−3D6580B/2022|EPI_ISL_12152094|2022−04−14
Scotland/QEUH−3A7E3D4/2022|EPI_ISL_11288673|2022−03−13
Scotland/QEUH−3A85282/2022|EPI_ISL_11286499|2022−03−13
0
5
10
15
20
C-to-T: 6, G-to-A: 4, T-to-C: 2, A-to-G: 1
Ireland/MH−NVRL−ecS22IRL00316051/2022|EPI_ISL_13775569|2022−05−02
Ireland/MH−NVRL−ecS22IRL00316053/2022|EPI_ISL_13775570|2022−05−02
NorthernIreland/NIRE−01c778/2022|EPI_ISL_12256802|2022−04−09
NorthernIreland/NIRE−01c429/2022|EPI_ISL_12102088|2022−03−30
Ireland/KE−NVRL−ecS22IRL00263443/2022|EPI_ISL_12585296|2022−03−22
Ireland/KE−NVRL−S22IRL00265211/2022|EPI_ISL_12316843|2022−03−22
Northern_Ireland/NIRE−01c5c0/2022|OX494349.1|2022−04−06
Northern_Ireland/NIRE−01c778/2022|OX503225.1|2022−04−09
NorthernIreland/NIRE−01c5c0/2022|EPI_ISL_12255500|2022−04−06
NorthernIreland/NIRE−01c8af/2022|EPI_ISL_12559305|2022−04−10
0
5
10

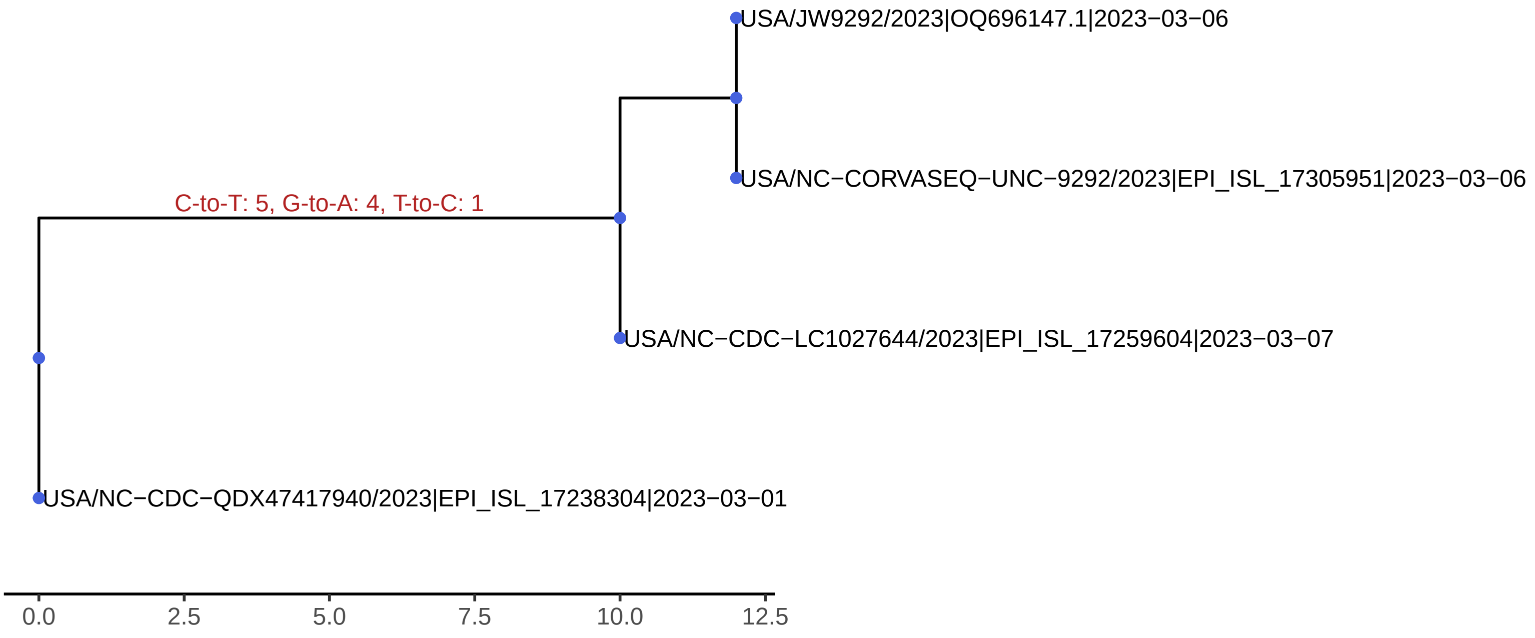
USA/JW9292/2023|OQ696147.1|2023−03−06
USA/NC−CORVASEQ−UNC−9292/2023|EPI_ISL_17305951|2023−03−06
C-to-T: 5, G-to-A: 4, T-to-C: 1
USA/NC−CDC−LC1027644/2023|EPI_ISL_17259604|2023−03−07
USA/NC−CDC−QDX47417940/2023|EPI_ISL_17238304|2023−03−01
0.0
2.5
5.0
7.5
10.0
12.5

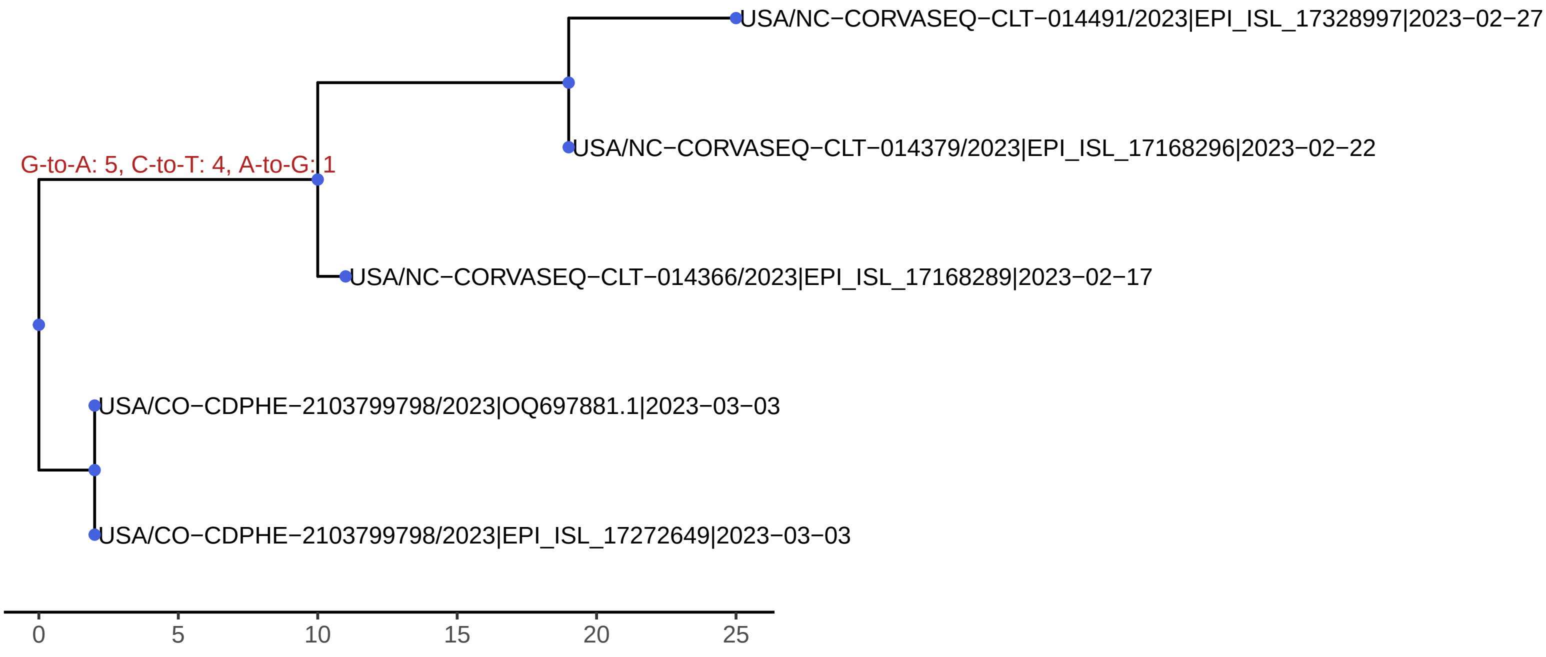
USA/NC−CORVASEQ−CLT−014491/2023|EPI_ISL_17328997|2023−02−27
USA/NC−CORVASEQ−CLT−014379/2023|EPI_ISL_17168296|2023−02−22
G-to-A: 5, C-to-T: 4, A-to-G: 1
USA/NC−CORVASEQ−CLT−014366/2023|EPI_ISL_17168289|2023−02−17
USA/CO−CDPHE−2103799798/2023|OQ697881.1|2023−03−03
USA/CO−CDPHE−2103799798/2023|EPI_ISL_17272649|2023−03−03
0
5
10
15
20
25

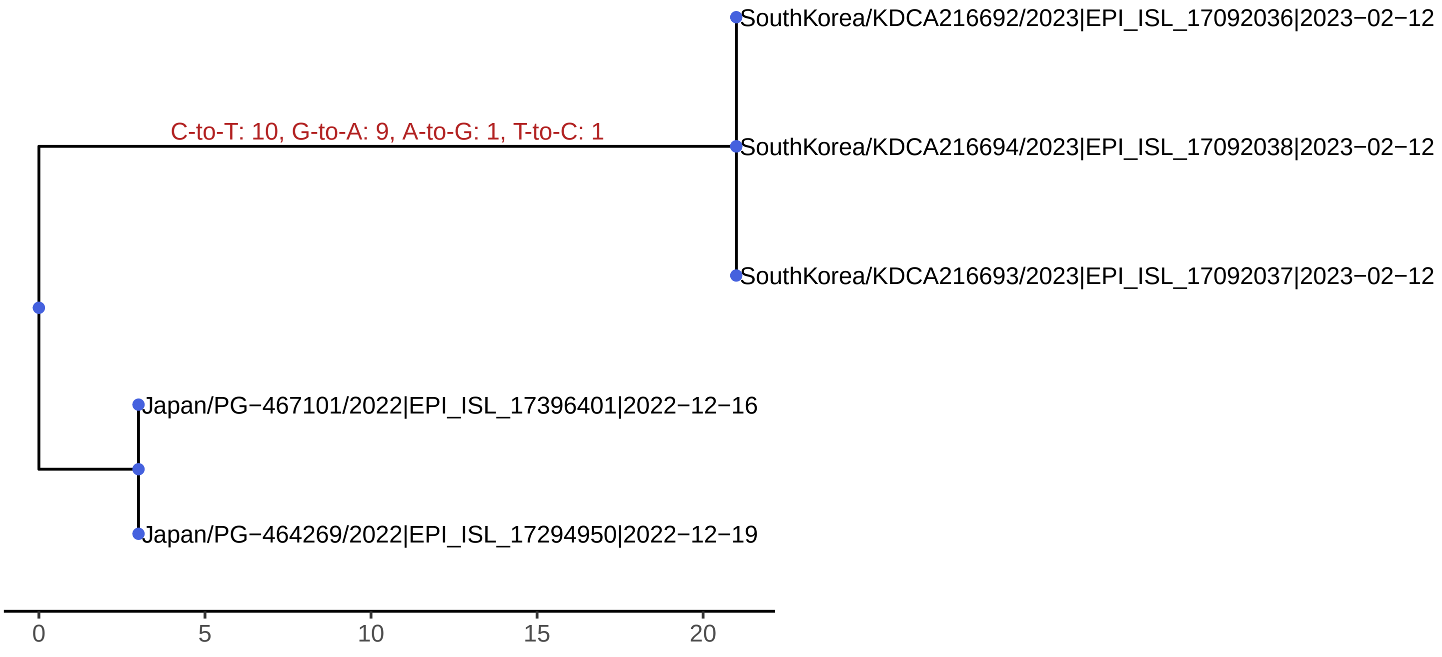
SouthKorea/KDCA216692/2023|EPI_ISL_17092036|2023−02−12
C-to-T: 10, G-to-A: 9, A-to-G: 1, T-to-C: 1
SouthKorea/KDCA216694/2023|EPI_ISL_17092038|2023−02−12
SouthKorea/KDCA216693/2023|EPI_ISL_17092037|2023−02−12
Japan/PG−467101/2022|EPI_ISL_17396401|2022−12−16
Japan/PG−464269/2022|EPI_ISL_17294950|2022−12−19
0
5
10
15
20

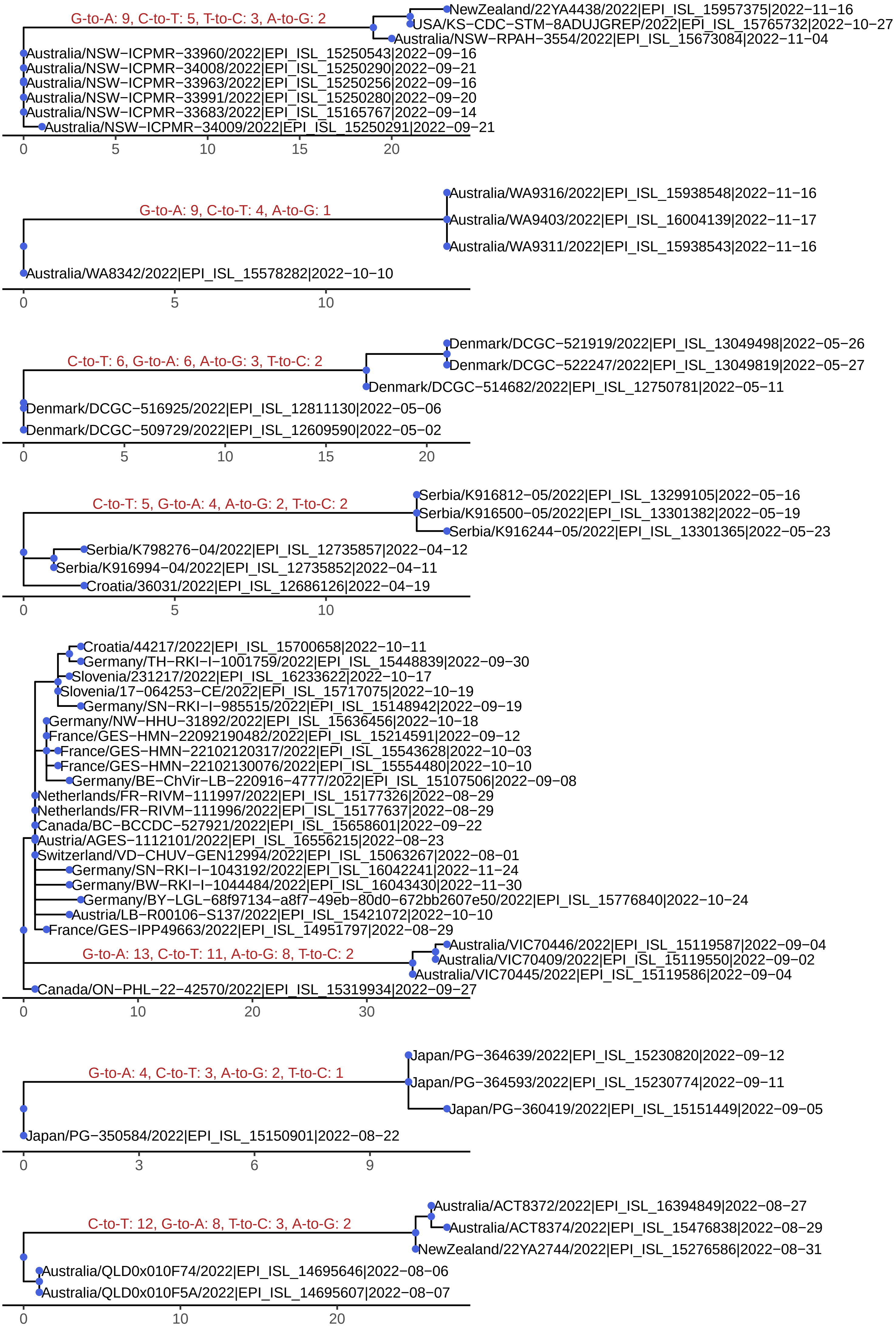

G-to-A: 9, C-to-T: 5, T-to-C: 3, A-to-G: 2
NewZealand/22YA4438/2022|EPI_ISL_15957375|2022−11−16
USA/KS−CDC−STM−8ADUJGREP/2022|EPI_ISL_15765732|2022−10−27
Australia/NSW−RPAH−3554/2022|EPI_ISL_15673084|2022−11−04
Australia/NSW−ICPMR−33960/2022|EPI_ISL_15250543|2022−09−16
Australia/NSW−ICPMR−34008/2022|EPI_ISL_15250290|2022−09−21
Australia/NSW−ICPMR−33963/2022|EPI_ISL_15250256|2022−09−16
Australia/NSW−ICPMR−33991/2022|EPI_ISL_15250280|2022−09−20
Australia/NSW−ICPMR−33683/2022|EPI_ISL_15165767|2022−09−14
Australia/NSW−ICPMR−34009/2022|EPI_ISL_15250291|2022−09−21
0
5
10
15
20
G-to-A: 9, C-to-T: 4, A-to-G: 1
Australia/WA9316/2022|EPI_ISL_15938548|2022−11−16
Australia/WA9403/2022|EPI_ISL_16004139|2022−11−17
Australia/WA9311/2022|EPI_ISL_15938543|2022−11−16
Australia/WA8342/2022|EPI_ISL_15578282|2022−10−10
0
5
10
C-to-T: 6, G-to-A: 6, A-to-G: 3, T-to-C: 2
Denmark/DCGC−521919/2022|EPI_ISL_13049498|2022−05−26
Denmark/DCGC−522247/2022|EPI_ISL_13049819|2022−05−27
Denmark/DCGC−514682/2022|EPI_ISL_12750781|2022−05−11
Denmark/DCGC−516925/2022|EPI_ISL_12811130|2022−05−06
Denmark/DCGC−509729/2022|EPI_ISL_12609590|2022−05−02
0
5
10
15
20
C-to-T: 5, G-to-A: 4, A-to-G: 2, T-to-C: 2
Serbia/K916812−05/2022|EPI_ISL_13299105|2022−05−16
Serbia/K916500−05/2022|EPI_ISL_13301382|2022−05−19
Serbia/K916244−05/2022|EPI_ISL_13301365|2022−05−23
Serbia/K798276−04/2022|EPI_ISL_12735857|2022−04−12
Serbia/K916994−04/2022|EPI_ISL_12735852|2022−04−11
Croatia/36031/2022|EPI_ISL_12686126|2022−04−19
0
5
10
Croatia/44217/2022|EPI_ISL_15700658|2022−10−11
Germany/TH−RKI−I−1001759/2022|EPI_ISL_15448839|2022−09−30
Slovenia/231217/2022|EPI_ISL_16233622|2022−10−17
Slovenia/17−064253−CE/2022|EPI_ISL_15717075|2022−10−19
Germany/SN−RKI−I−985515/2022|EPI_ISL_15148942|2022−09−19
Germany/NW−HHU−31892/2022|EPI_ISL_15636456|2022−10−18
France/GES−HMN−22092190482/2022|EPI_ISL_15214591|2022−09−12
France/GES−HMN−22102120317/2022|EPI_ISL_15543628|2022−10−03
France/GES−HMN−22102130076/2022|EPI_ISL_15554480|2022−10−10
Germany/BE−ChVir−LB−220916−4777/2022|EPI_ISL_15107506|2022−09−08
Netherlands/FR−RIVM−111997/2022|EPI_ISL_15177326|2022−08−29
Netherlands/FR−RIVM−111996/2022|EPI_ISL_15177637|2022−08−29
Canada/BC−BCCDC−527921/2022|EPI_ISL_15658601|2022−09−22
Austria/AGES−1112101/2022|EPI_ISL_16556215|2022−08−23
Switzerland/VD−CHUV−GEN12994/2022|EPI_ISL_15063267|2022−08−01
Germany/SN−RKI−I−1043192/2022|EPI_ISL_16042241|2022−11−24
Germany/BW−RKI−I−1044484/2022|EPI_ISL_16043430|2022−11−30
Germany/BY−LGL−68f97134−a8f7−49eb−80d0−672bb2607e50/2022|EPI_ISL_15776840|2022−10−24
Austria/LB−R00106−S137/2022|EPI_ISL_15421072|2022−10−10
France/GES−IPP49663/2022|EPI_ISL_14951797|2022−08−29
G-to-A: 13, C-to-T: 11, A-to-G: 8, T-to-C: 2
Australia/VIC70446/2022|EPI_ISL_15119587|2022−09−04
Australia/VIC70409/2022|EPI_ISL_15119550|2022−09−02
Australia/VIC70445/2022|EPI_ISL_15119586|2022−09−04
Canada/ON−PHL−22−42570/2022|EPI_ISL_15319934|2022−09−27
0
10
20
30
G-to-A: 4, C-to-T: 3, A-to-G: 2, T-to-C: 1
Japan/PG−364639/2022|EPI_ISL_15230820|2022−09−12
Japan/PG−364593/2022|EPI_ISL_15230774|2022−09−11
Japan/PG−360419/2022|EPI_ISL_15151449|2022−09−05
Japan/PG−350584/2022|EPI_ISL_15150901|2022−08−22
0
3
6
9
C-to-T: 12, G-to-A: 8, T-to-C: 3, A-to-G: 2
Australia/ACT8372/2022|EPI_ISL_16394849|2022−08−27
Australia/ACT8374/2022|EPI_ISL_15476838|2022−08−29
NewZealand/22YA2744/2022|EPI_ISL_15276586|2022−08−31
Australia/QLD0x010F74/2022|EPI_ISL_14695646|2022−08−06
Australia/QLD0x010F5A/2022|EPI_ISL_14695607|2022−08−07
0
10
20
